# Psychological factors associated with reporting side effects following COVID-19 vaccination: a prospective cohort study (CoVAccS – wave 3)

**DOI:** 10.1101/2022.09.07.22279682

**Authors:** Louise E. Smith, Julius Sim, Susan M. Sherman, Richard Amlôt, Megan Cutts, Hannah Dasch, Nick Sevdalis, G James Rubin

## Abstract

**Objective:** To investigate symptom reporting following the first and second COVID-19 vaccine doses, attribution of symptoms to the vaccine, and factors associated with symptom reporting.

**Methods:** Prospective cohort study (T1: 13-15 January 2021, T2: 4-15 October 2021). Participants were aged 18 years or older, living in the UK. Personal, clinical, and psychological factors were investigated at T1. Symptoms were reported at T2. We used logistic regression analyses to investigate associations.

**Results:** After the first COVID-19 vaccine dose, 74.1% (95% CI 71.4% to 76.7%, *n*=762/1028) of participants reported at least one injection-site symptom, while 65.0% (95% CI 62.0% to 67.9%, *n*=669/1029) reported at least one other (non-injection-site) symptom. Symptom reporting was associated with being a woman and younger. After the second dose, 52.9% (95% CI 49.8% to 56.0%, *n*=532/1005) of participants reported at least one injection-site symptom and 43.7% (95% CI 40.7% to 46.8%, *n*=440/1006) reported at least one other (non-injection-site) symptom. Symptom reporting was associated with having reported symptoms after the first dose, having an illness that put one at higher risk of COVID-19 (non-injection-site symptoms only), and not believing that one had enough information about COVID-19 to make an informed decision about vaccination (injection-site symptoms only).

**Conclusions:** Women and younger people were more likely to report symptoms from vaccination. People who had reported symptoms from previous doses were also more likely to report symptoms subsequently, although symptom reporting following the second vaccine was lower than following the first vaccine. Few psychological factors were associated with symptom reporting.

**Highlights:**

- We measured symptom reporting and attributions from the COVID-19 vaccines.
- A prospective cohort study was used (T1: January 2021, T2: October 2021).
- Women and younger people were more likely to report side effects.
- Side effects reporting after the first and second dose was strongly associated.

## Introduction

Side effects can occur after taking a medication, including being vaccinated. While some side effects may be due to the pharmacological action of the drug, others may arise from the so-called ‘nocebo effect’, a phenomenon whereby the expectation that symptoms will develop becomes self-fulfilling.^1^ The role of expectation in later perception of side effects has been well-documented for medications^2 3^ and vaccinations.^4^ Psychosocial factors may contribute to the nocebo effect; for example, seeing or hearing that a vaccine causes side effects,^5^ or holding more negative beliefs about medications.^6 7^

Clinical trial data for UK approved COVID-19 vaccines (AstraZeneca, Pfizer-BioNTech, Moderna) indicate that side-effect reporting is lower in older adults.^8-10^ However, the association with vaccine dose is less clear cut, with different patterns emerging for the different vaccines. For the AstraZeneca vaccine, side-effect reporting was lower after the second dose than after the first.^8^ For the Pfizer-BioNTech vaccine, there was no difference in the percentage of people experiencing local reactions following the first and second dose.^9 11^ One UK study investigating symptom reporting on an app (over 627,000 vaccinated app users) found that people who had the Pfizer-BioNTech vaccine reported more systemic effects (e.g. fatigue and headache) after the second dose, compared to the first.^12^ The pattern for reporting systemic reactions differed by previous SARS-CoV-2 infection, with those with evidence of previous infection reporting more systemic reactions after the first dose and those with no evidence of previous infection reporting more systemic reactions after the second dose.^11^ Reported adverse effects for the Moderna vaccine were more severe following the second than following the first dose.^10^

Research investigating symptom reporting following vaccination for COVID-19 has focused on the socio-demographic factors associated with symptom reporting. One US survey (over 19,000 respondents) found that reporting adverse effects was associated with being younger, female, Asian ethnicity (compared to white), having had COVID-19 before, and it being the second vaccine dose (Moderna and Pfizer-BioNTech).^13^ A study of UK app users (over 627,000 respondents) found that women, younger people, and those with previous SARS-CoV-2 infection were more likely to report symptoms (local and systemic, both AstraZeneca and Pfizer-BioNTech), but found no clear trend with vaccine type, dose or comorbidity.^12^

Fewer studies have investigated the association between psychological factors and COVID-19 vaccine side-effect reporting. Where they have, studies have focused on the influence of seeing or hearing about symptom reporting in others. For example, one study found that seeing more social media posts about COVID-19 vaccine side effects and severity of impressions from news stories and personal contacts were associated with later experiencing side effects.^14^ Another study found that following the reporting of severe adverse events in the media, reporting to the national Centre for Adverse Reactions Monitoring (New Zealand) for effects mentioned in the media increased, whereas there was no change in the reporting of adverse events that were not specifically mentioned.^15^ One study, investigating other psychological variables, found evidence for an association between COVID-19 vaccine side-effect reports and higher side-effect expectations, greater worry about COVID-19, and depressive symptoms.^16^

At the start of the COVID-19 vaccine rollout in the UK (January 2021), we conducted an online cross-sectional survey investigating perceptions about COVID-19 and vaccination, vaccination intention and side-effect expectations.^17^ We found that only 9% of participants thought that side effects were likely (58% judged them uncertain, 33% judged them unlikely), while clinical trial data indicated that rates experienced were substantially higher (injection-site symptoms up to 89%, non-injection-site symptoms up to 70%). Higher expectations that one would experience side effects from a COVID-19 vaccine were associated with older age, being clinically extremely vulnerable to COVID-19, being afraid of needles, perceiving lower social norms for COVID-19 vaccination, lower perceived necessity and safety of COVID-19 vaccination, and not thinking that one had enough information about COVID-19 vaccination or illness.

In this study we used results from a follow-up survey, conducted in October 2021 after all UK adults had been offered two doses of the vaccine,^18^ to investigate prevalence of injection- and non-injection-site symptoms following COVID-19 vaccination and their attribution to the vaccine. We investigated associations between symptom reporting (injection- and non-injection-site) and personal, clinical, psychological, and contextual factors following the first and second COVID-19 vaccine doses separately.

## Methods

### Design

Prospective cohort study conducted at two timepoints (T1, 13–15 January 2021, *n*=1500; T2, 4–15 October 2021, *n*=1148, response rate 76.5%). For more details on the study design, see Smith et al.^18^

### Participants

People were eligible for the study if they lived in the UK, were aged 18 years or older, and had not completed our previous survey (due to similarities in questionnaire materials).^19^ Participants were recruited to the study from Prolific’s online research panel (people who have signed up to take part in online surveys). Quota sampling was used, based on age, sex, and ethnicity, so that participant characteristics in these respects were similar to those of the UK population. Consent was provided before starting the survey. Participants were paid £2 per survey upon completion.

For this study, we excluded people who reported having been vaccinated for COVID-19 at T1 (*n*=30). As the outcome measures are symptom reporting following vaccination, only participants who reported that they had been vaccinated for COVID-19 (one or two doses) were selected (first vaccine dose, *n*=1034; second vaccine dose, *n*=1009).

### Measures

Survey materials are available online.^20 21^

#### Outcome measures

We measured symptom reporting and attribution to the vaccine at T2 using items based on the Side Effect Attribution Scale.^22^ Participants were asked if they experienced any symptoms (from a list of thirteen: seven injection-site symptoms, six non-injection-site symptoms; symptoms based on Menni et al.^12^) in the seven days after they received a COVID-19 vaccine. We asked participants about each symptom on a six-point scale (“no”, “yes, but definitely not a side effect”, “yes, but probably not a side effect”, “yes, but unsure whether a side effect”, “yes, and probably a side effect” and “yes, and definitely a side effect”). Participants were categorised as having injection-site symptoms if they reported experiencing any of the seven injection-site symptoms after vaccination. Participants were categorised as having non-injection-site symptoms if they reported experiencing any of the six non-injection-site symptoms after vaccination. We asked about participants’ first and second dose of the vaccine separately.

#### Personal and clinical factors

Participants were asked for their age, gender, ethnicity, and whether they thought they had previously had COVID-19 or currently had it at T1. Answers were recoded into a binary variable (“definitely” and “probably” had it or have it now vs “definitely” and “probably” not had it and do not have it now; “don’t know” and “prefer not to say” were coded as missing). At T2, participants were asked whether they had a chronic illness. We recoded this into a binary variable indicating whether the participant was at high risk for COVID-19 or not.

#### Psychological and contextual factors

At T1, participants were asked about their COVID-19 vaccination beliefs and attitudes using a series of seventeen items rated on an eleven-point (0–10) scale anchored at ‘strongly disagree’ (0) and ‘strongly agree’ (10). In previous analyses, these items were subjected to a principal components analysis, resulting in five components.^23^ Participants were also asked whether they were afraid of needles on the same 0–10 scale.

Side-effect expectations were measured at T1 with a single item asking how likely participants thought they were to get side effects from a COVID-19 vaccine on a 0–10 scale anchored at ‘extremely unlikely’ (0) and ‘extremely likely’ (10).

### Ethics

Ethical approval for this study was granted by Keele University’s Research Ethics Committee (reference: PS-200129).

### Power

We had more than 99% power to detect a small odds ratio (1.53, in respect of a binary predictor),^24^ with a sample size of 1005, α=0.05, and an assumed outcome prevalence of 40%.

### Analysis

Responses to side-effect questions were not forced; there was, therefore, a small amount of missing data for individual symptoms (first vaccine dose, up to 0.6%, *n*=6/1034; second vaccine dose, up to 0.4%, *n*=4/1009).

Reporting and attribution of symptoms was investigated following the first and second COVID-19 vaccine doses separately.

We recoded symptom reporting into a binary variable (symptom not reported, vs one or more symptom reported). For reported symptoms, we also recoded symptom attribution into a binary variable (definitely not, probably not, vs unsure, probably, definitely). We categorised symptoms into two categories: i) injection-site symptoms (pain or tenderness where the injection was, redness where the injection was, swelling where the injection was, itch where the injection was, warmth where the injection was, bruising where the injection was, other symptom[s] where the injection was), and ii) non-injection-site symptoms (diarrhoea, headache, joint or muscle pain, high temperature/fever, nausea, fatigue). For each vaccine dose, we then created two further variables indicating whether the participant had experienced any injection-site symptoms (no injection-site symptoms reported, vs injection-site symptom reported) or any non-injection-site symptoms (no non-injection-site symptom reported, vs non-injection-site symptom reported) respectively. We did the same for attribution (of symptoms to vaccine).

Symptom reporting and attribution of individual symptoms, and injection- and non-injection-site symptoms, are reported descriptively. A chi-squared test was used to investigate whether there was an association between reporting injection-site symptoms and non-injection-site symptoms.

We investigated factors associated with reporting of injection-site and non-injection-site symptoms separately, using binary logistic regression. Explanatory variables (except for at risk for COVID-19) were measured at T1, while the outcomes (symptom reporting) were reported at T2. Explanatory variables were entered into the logistic regression model in two blocks, selected *a priori* based on previous analyses.^18 23^ We entered personal and clinical characteristics into the first block: age, gender, ethnicity, at risk for COVID-19, think or had COVID-19 previously or currently, and vaccine brand. Psychological and contextual factors were entered into the second block: fear of needles, four principal vaccine components (social norms relating to vaccination, perceived necessity of vaccination, perceived safety of the vaccine, adequacy of information about the vaccine) and side-effect expectations. In analyses investigating symptom reporting following the second vaccine dose we also included a single item in a third block: symptom reporting following the first vaccine. The Nagelkerke (pseudo-) *R*^2^ was used to investigate the predictive strength of the regression models; this statistic can take values between 0 and 1.

Statistical significance was set at *p* ≤.05.

We did not conduct analyses of associations with symptom attribution owing to the very small number of symptoms that were not attributed to vaccination, in relation to both the first and the second vaccine.

## Results

### Participant characteristics

After excluding those who had been vaccinated at T1, 89.8% (*n*=1009/1124) reported having received two vaccine doses, with 2.2% (*n*=25) reporting having had one vaccine only (7.6%, *n*=85 not vaccinated; 0.4%, *n*=4 preferred not to say; 0.1%, *n*=1 did not know).

Of those who had been vaccinated at T2, 52.1% were female (*n*=539/1034; 47.6% male, *n*=492; 0.2% non-binary, *n*=2; 0.1% prefer to self-describe, *n*=1), 86.9% were white (*n*=899/1034; 12.7% Black or minority ethnicities, *n*=131; 0.4% prefer not to say, *n*=4), with a mean age of 48.7 years (SD=15.1, range 18 to 80 years).

The most commonly reported vaccine received was AstraZeneca (57.7%, *n*=597/1034) followed by: Pfizer-BioNTech (38.2%, *n*=395); Moderna (3.5%, *n*=36); Janssen (0.1%, *n*=1); a vaccine not listed, (0.3% *n*=3); did not know (0.2%, *n*=2).

### Symptom reporting and attribution to COVID-19 vaccination

The most common injection-site symptom reported following the first and second doses of a COVID-19 vaccine was pain or tenderness where the injection was (Table 1). The most common non-injection-site symptoms were fatigue, joint or muscle pain, and headache.

**Table 1.**
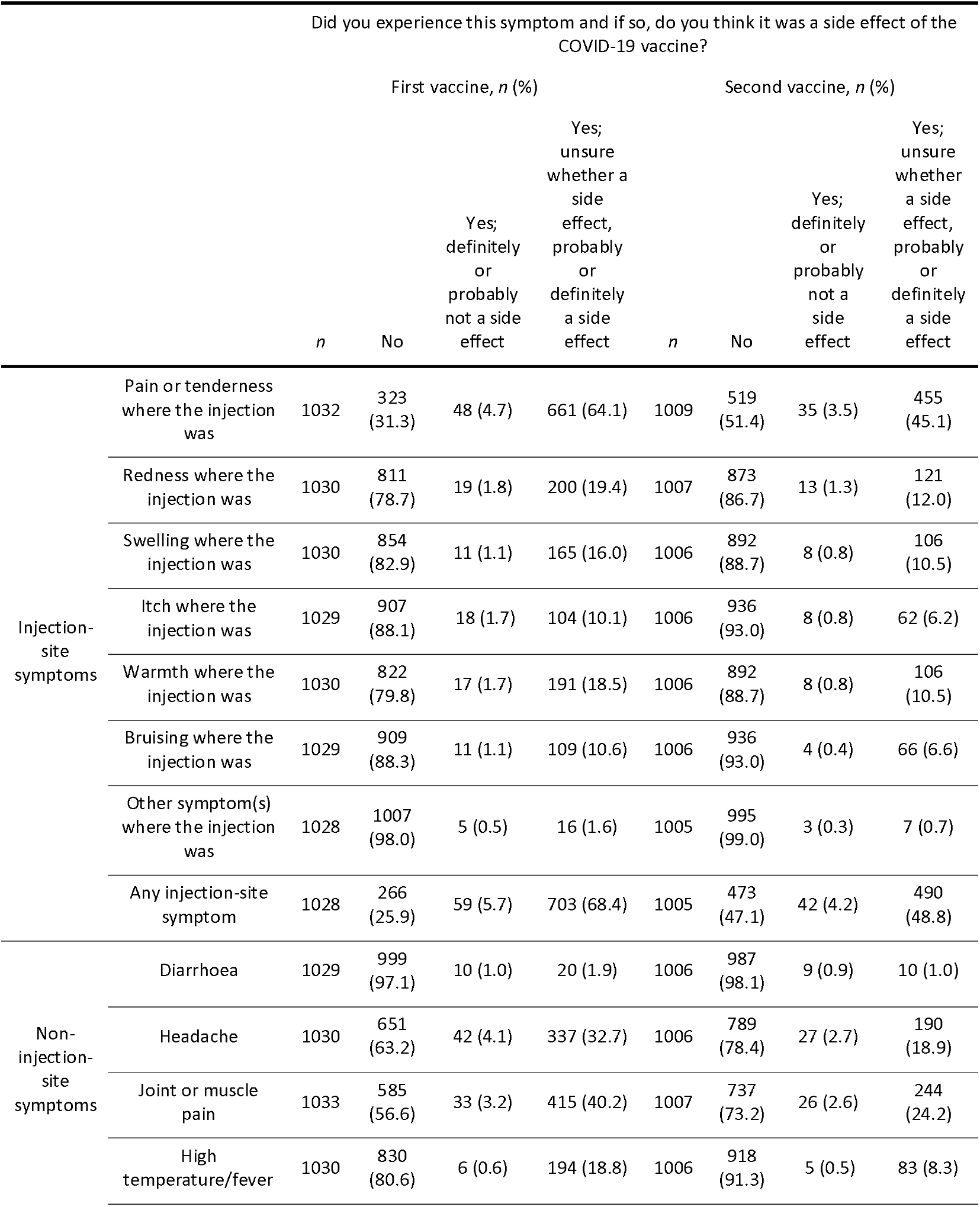
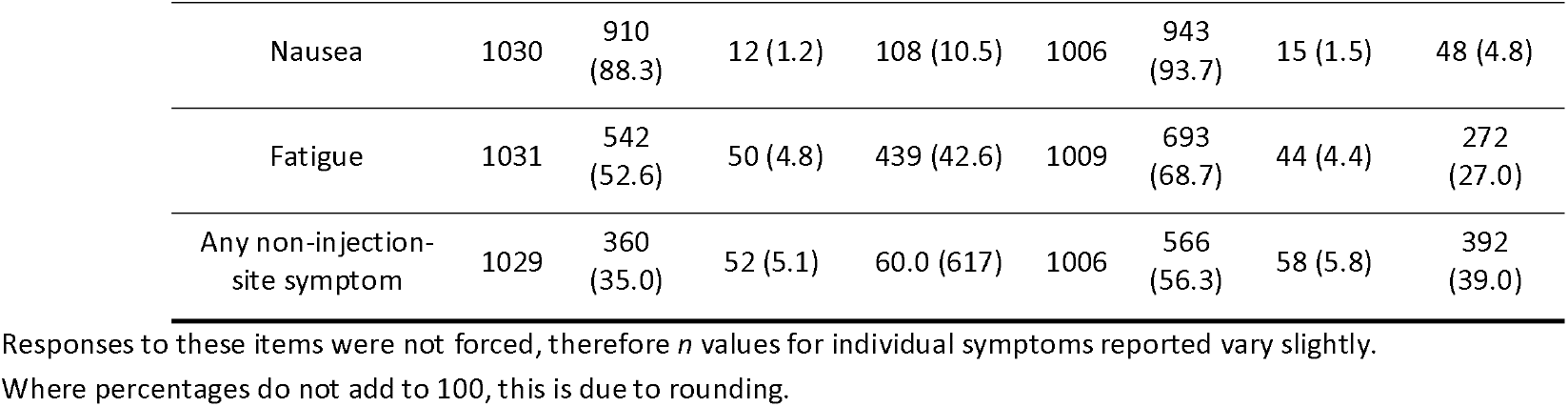
Symptom reporting and attribution to COVID-19 vaccination for the first and second vaccine dose separately.

|  |  | Did you experience this symptom and if so, do you think it was a side effect of the COVID-19 vaccine? |  |  |  |  |  |  |  |
| --- | --- | --- | --- | --- | --- | --- | --- | --- | --- |
|  |  | First vaccine, *n* (%) |  |  |  | Second vaccine, *n* (%) |  |  |  |
|  |  | *n* | No | Yes; definitely or probably not a side effect | Yes; unsure whether a side effect, probably or definitely a side effect | *n* | No | Yes; definitely or probably not a side effect | Yes; unsure whether a side effect, probably or definitely a side effect |
| Injection-site symptoms | Pain or tenderness where the injection was | 1032 | 323 (31.3) | 48 (4.7) | 661 (64.1) | 1009 | 519 (51.4) | 35 (3.5) | 455 (45.1) |
|  | Redness where the injection was | 1030 | 811 (78.7) | 19 (1.8) | 200 (19.4) | 1007 | 873 (86.7) | 13 (1.3) | 121 (12.0) |
|  | Swelling where the injection was | 1030 | 854 (82.9) | 11 (1.1) | 165 (16.0) | 1006 | 892 (88.7) | 8 (0.8) | 106 (10.5) |
|  | Itch where the injection was | 1029 | 907 (88.1) | 18 (1.7) | 104 (10.1) | 1006 | 936 (93.0) | 8 (0.8) | 62 (6.2) |
|  | Warmth where the injection was | 1030 | 822 (79.8) | 17 (1.7) | 191 (18.5) | 1006 | 892 (88.7) | 8 (0.8) | 106 (10.5) |
|  | Bruising where the injection was | 1029 | 909 (88.3) | 11 (1.1) | 109 (10.6) | 1006 | 936 (93.0) | 4 (0.4) | 66 (6.6) |
|  | Other symptom(s) where the injection was | 1028 | 1007 (98.0) | 5 (0.5) | 16 (1.6) | 1005 | 995 (99.0) | 3 (0.3) | 7 (0.7) |
| Any injection-site symptom |  | 1028 | 266 (25.9) | 59 (5.7) | 703 (68.4) | 1005 | 473 (47.1) | 42 (4.2) | 490 (48.8) |
| Non-injection-site symptoms | Diarrhoea | 1029 | 999 (97.1) | 10 (1.0) | 20 (1.9) | 1006 | 987 (98.1) | 9 (0.9) | 10 (1.0) |
|  | Headache | 1030 | 651 (63.2) | 42 (4.1) | 337 (32.7) | 1006 | 789 (78.4) | 27 (2.7) | 190 (18.9) |
|  | Joint or muscle pain | 1033 | 585 (56.6) | 33 (3.2) | 415 (40.2) | 1007 | 737 (73.2) | 26 (2.6) | 244 (24.2) |
|  | High temperature/fever | 1030 | 830 (80.6) | 6 (0.6) | 194 (18.8) | 1006 | 918 (91.3) | 5 (0.5) | 83 (8.3) |

|  |  |  |  |  |  |  |  |  |
| --- | --- | --- | --- | --- | --- | --- | --- | --- |
| Nausea | 1030 | 910<br>(88.3) | 12 (1.2) | 108 (10.5) | 1006 | 943<br>(93.7) | 15 (1.5) | 48 (4.8) |
| Fatigue | 1031 | 542<br>(52.6) | 50 (4.8) | 439 (42.6) | 1009 | 693<br>(68.7) | 44 (4.4) | 272<br>(27.0) |
| Any non-injection-site symptom | 1029 | 360<br>(35.0) | 52 (5.1) | 60.0 (617) | 1006 | 566<br>(56.3) | 58 (5.8) | 392<br>(39.0) |

Responses to these items were not forced, therefore *n* values for individual symptoms reported vary slightly. Where percentages do not add to 100, this is due to rounding.

Following the first dose of the vaccine, 74.1% (95% CI 71.4% to 76.7%, *n*=762/1028) of participants reported experiencing at least one injection-site symptom. Of these, 92.3% (95% CI 90.1% to 94.0%, *n*=703/762) attributed at least one of symptom experienced to the vaccine. 65.0% (95% CI 62.0% to 67.9%, *n*=669/1029) reported experiencing at least one other (non-injection-site) symptom. Of these, 92.2% (95% CI 89.9% to 94.0%, *n*=617/669) attributed at least one of symptom experienced to the vaccine.

Following the second dose of the vaccine, 52.9% (95% CI 49.8% to 56.0%, *n*=532/1005) of participants reported experiencing at least one injection-site symptom. Of these, 92.1% (95% CI 89. 5% to 94.1%, *n*=490/532) attributed at least one of symptom experienced to the vaccine. 43.7% (95% CI 40.7% to 46.8%, *n*=440/1006) reported experiencing at least one other (non-injection-site) symptom. Of these, 89.1% (95% CI 85.8% to 91.7%, *n*=392/440) attributed at least one symptom experienced to the vaccine.

There was a significant difference in reporting of injection-site and non-injection-site symptoms for both vaccines, with 354/1028 (34.4%) of participants reporting one type of symptom but not the other for the first vaccine (*χ*^2^_1_ = 40.9, *p*<0.001) and a corresponding figure of 367/1005 (36.5%) for the second vaccine dose (*χ*^2^_1_ = 78.7, *p*<0.001; Table 2).

**Table 2.** Number of people reporting injection- and non-injection-site symptoms after the first and second vaccine dose.

|  |  | First vaccine, *n*=1028 |  | Second vaccine, *n*=1005 |  |
| --- | --- | --- | --- | --- | --- |
|  |  | Non-injection-site symptoms, *n* (%) |  | Non-injection-site symptoms, *n* (%) |  |
|  |  | Not reported | Reported | Not reported | Reported |
| Injection-site symptoms | Not reported | 136 (37.8) | 130 (19.5) | 336 (59.4) | 137 (31.2) |
|  | Reported | 224 (62.2) | 538 (80.5) | 230 (40.6) | 302 (68.8) |
| Total |  | 360 (100.0) | 668 (100) | 566 (100) | 439 (100) |

### Associations between symptom reporting and personal, clinical, psychological, and contextual factors

Results reported are from the full regression model. Results from block 1 alone are presented in Appendix A. Descriptive statistics relating to variables in the regression models are presented in Table 3. Missing data in regression models are due to missing values for individual variables.

**Table 3.** Participant characteristics, in subgroups according to dose and symptom reporting. Data are n (%) except where indicated otherwise

|  |  | First vaccine, *n* (%) |  |  |  | Second vaccine, *n* (%) |  |  |  |
| --- | --- | --- | --- | --- | --- | --- | --- | --- | --- |
|  |  | Injection-site symptoms |  | Non-injection-site symptoms |  | Injection-site symptoms |  | Non-injection-site symptoms |  |
|  |  | Not reported<br>*n*=234 | Reported<br>*n*=666 | Not reported<br>*n*=234 | Reported<br>*n*=666 | Not reported<br>*n*=419 | Reported<br>*n*=463 | Not reported<br>*n*=501 | Reported<br>*n*=382 |
| Age (years); mean (SD) |  | 53.4 (14.4) | 47.1 (15.2) | 52.7 (14.9) | 46.6 (15.0) | 52.0 (14.4) | 46.3 (15.3) | 51.6 (14.1) | 45.5 (15.7) |
| Gender | Female | 82 (17.5) | 386 (82.5) | 143 (30.5) | 326 (69.5) | 192 (41.6) | 269 (58.4) | 242 (52.4) | 220 (47.6) |
|  | Male | 152 (35.2) | 280 (64.8) | 170 (39.4) | 262 (60.6) | 227 (53.9) | 194 (46.1) | 259 (61.5) | 162 (38.5) |
| Ethnicity | White | 209 (26.4) | 582 (73.6) | 284 (35.9) | 508 (64.1) | 377 (48.7) | 397 (51.3) | 451 (58.2) | 324 (41.8) |
|  | Other ethnic groups | 25 (22.9) | 84 (77.1) | 29 (26.6) | 80 (73.4) | 42 (38.9) | 66 (61.1) | 50 (46.3) | 58 (53.7) |
| At risk for COVID-19 | No | 187 (25.4) | 548 (74.6) | 244 (33.2) | 491 (66.8) | 341 (47.4) | 379 (52.6) | 412 (57.2) | 308 (42.8) |
|  | Yes | 47 (28.5) | 118 (71.5) | 69 (41.6) | 97 (58.4) | 78 (48.1) | 84 (51.9) | 89 (54.6) | 74 (45.4) |
| Think previously or currently had COVID-19 | No | 205 (26.8) | 559 (73.2) | 277 (36.3) | 487 (63.7) | 369 (49.1) | 383 (50.9) | 444 (59.0) | 308 (41.0) |
|  | Yes | 29 (21.3) | 107 (78.7) | 36 (26.3) | 101 (73.7) | 50 (38.5) | 80 (61.5) | 57 (43.5) | 74 (56.5) |
| Vaccine brand | Pfizer-BioNTech | 64 (17.9) | 293 (82.1) | 142 (39.8) | 215 (60.2) | 128 (36.9) | 219 (63.1) | 176 (50.7) | 171 (49.3) |
|  | AstraZeneca | 168 (32.4) | 350 (67.6) | 164 (31.6) | 355 (68.4) | 287 (56.1) | 225 (43.9) | 322 (62.8) | 191 (37.2) |
|  | Moderna | 2 (8.0) | 23 (92.0) | 7 (28.0) | 18 (72.0) | 4 (17.4) | 19 (82.6) | 3 (13.0) | 20 (87.0) |
| I am afraid of needles (0–10, strongly disagree to strongly agree); mean (SD) |  | 2.2 (3.1) | 2.8 (3.3) | 2.3 (3.2) | 2.8 (3.3) | 2.4 (3.2) | 2.8 (3.4) | 2.5 (3.3) | 2.8 (3.3) |
| How likely to get side effects from vaccine? (0–10, strongly disagree to strongly agree); mean (SD) |  | 3.6 (2.4) | 3.7 (2.3) | 3.6 (2.4) | 3.7 (2.3) | 3.5 (2.3) | 3.8 (2.3) | 3.5 (2.3) | 3.8 (2.3) |
| Symptoms reported after first vaccine dose | No | — | — | — | — | 207 (89.2) | 25 (10.8) | 266 (86.6) | 41 (13.4) |
|  | Yes | — | — | — | — | 212 (32.6) | 438 (67.4) | 235 (40.8) | 341 (59.2) |

SD = standard deviation

#### First vaccine dose

After the first vaccine dose, reporting injection-site and non-injection-site symptoms was associated with being female and younger (Table 4). Reporting symptoms at either site was also associated with vaccine brand. Examination of Table 3 reveals that reporting was highest for the Moderna vaccine, at 92% and 72% for vaccine-site and non-vaccine-site symptoms respectively, though these estimates are imprecise in view of the small number of cases in this category. Reporting was somewhat higher for the Pfizer-BioNTech vaccine than for the AstraZeneca vaccine for injection-site symptoms, but the reverse was the case for non-injection-site symptoms.

**Table 4.**
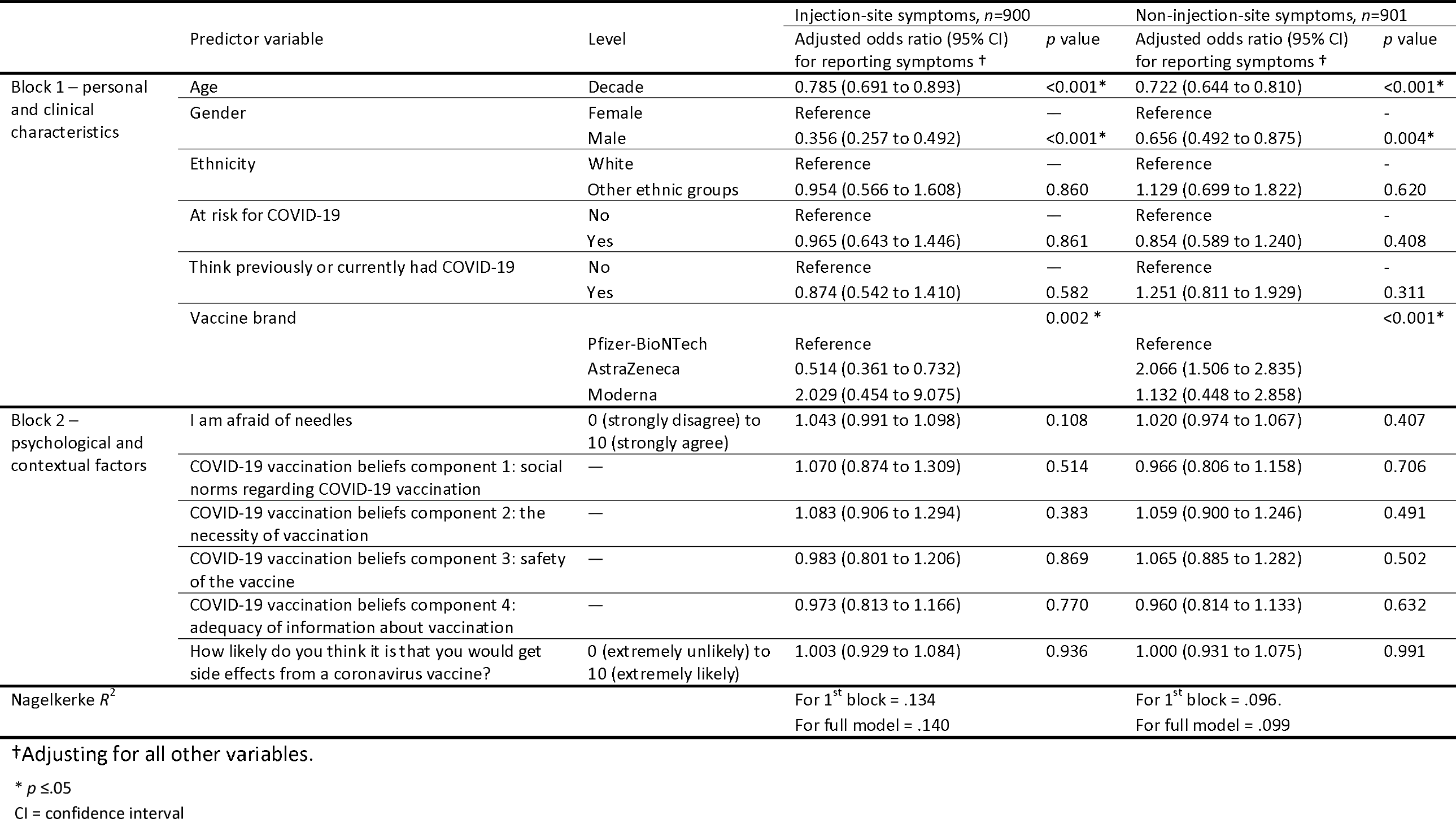
Results of the full logistic regression models analysing associations with symptom reporting following the first dose of a COVID-19 vaccination. Parameter estimates relate to the full model containing all explanatory variables (injection-site symptoms, n=900, 13.0% missing data; non-injection-site symptoms, n=901, 12.9% missing data). For continuous variables, the adjusted odds ratios represent the change in likelihood of side effects for a one-unit increase in the predictor variable, apart from age, where an increase of one-unit represents an increase by decade.

The regression model for injection-site symptoms had greater predictive power (Nagelkerke *R*^2^ = .140) than that for non-injection-site-symptoms (Nagelkerke *R*^2^ = .099). In both cases, the addition of psychological and contextual factors in the second block produced only a small increase in the *R*^2^ value over that derived from the personal and clinical variables in the first block.

#### Second vaccine dose

In respect of both injection-site and non-injection-site symptoms, those having reported symptoms after the first dose were much more likely to do so after the second dose (Table 5). Reporting injection- and non-injection-site symptoms after the second vaccine dose was associated with vaccine brand, in relation to both types of symptoms. As in the case of first-dose symptoms, the highest rate reported was for the Moderna vaccine (82.6% and 87.0% for injection-site and non-injection-site symptoms, respectively; Table 3). The rate was higher for the Pfizer-BioNTech than for the AstraZeneca vaccine in both cases. Reporting non-injection-site symptoms was associated with having an illness that put one at higher risk of COVID-19. Reporting injection-site symptoms was associated with not believing that one had enough information about COVID-19 (illness and vaccination) to make an informed decision about vaccination.

**Table 5.**
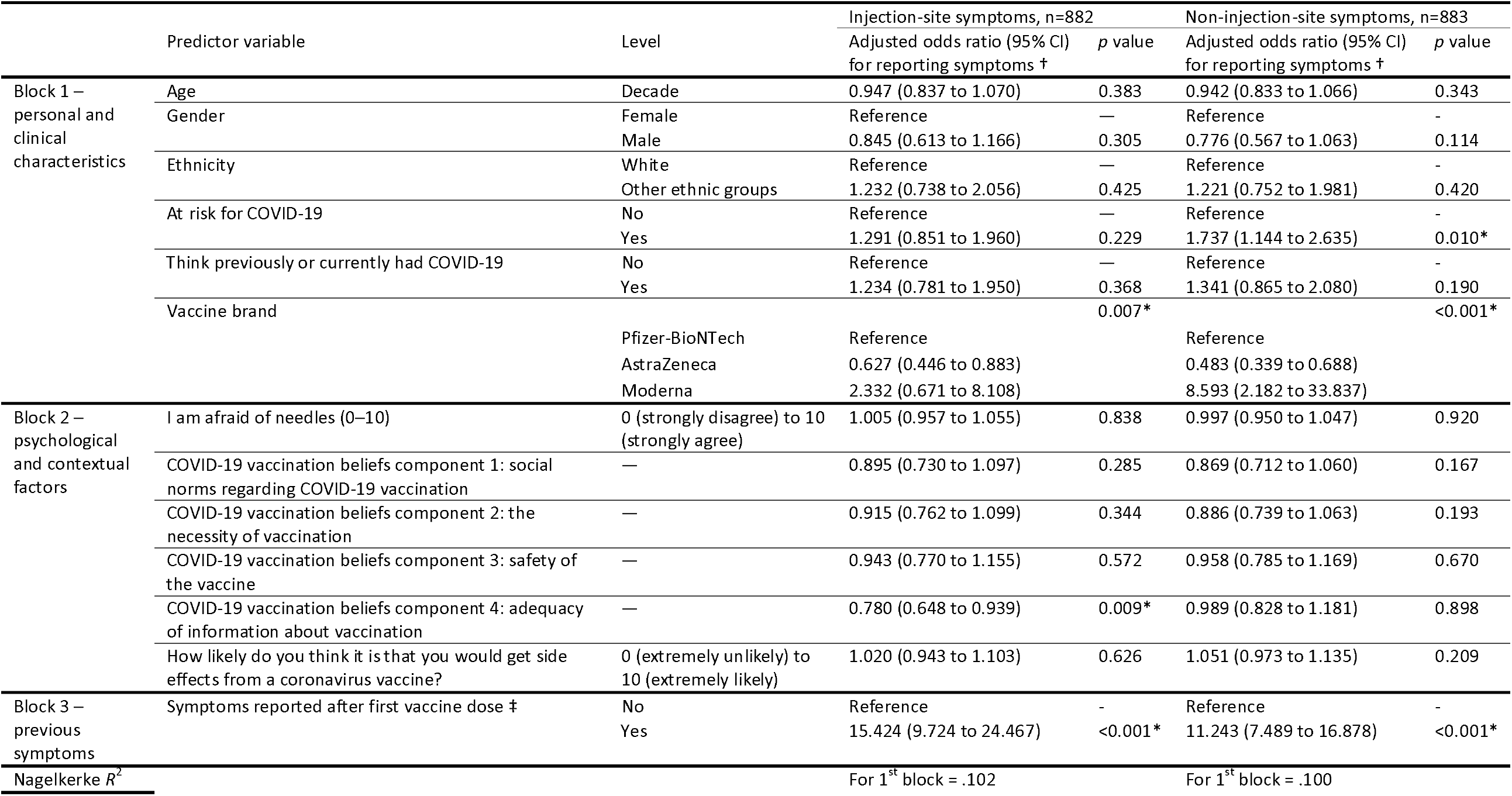
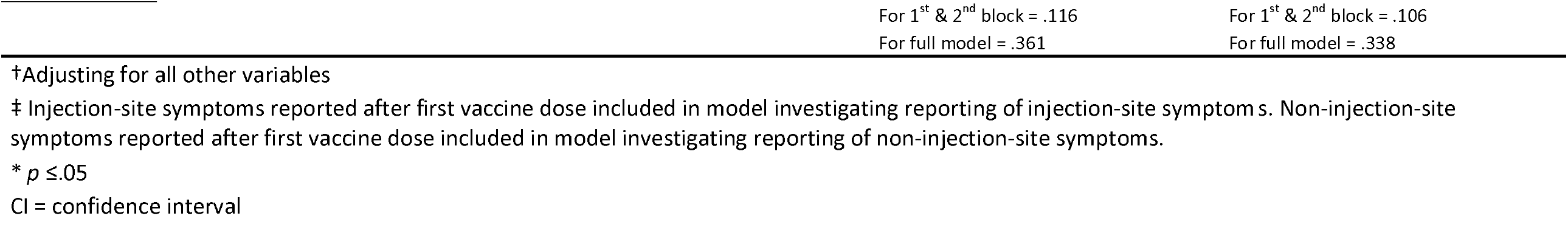
Results of the full logistic regression models analysing associations with symptom reporting following the second dose of a COVID-19 vaccination. Parameter estimates relate to the full model containing all explanatory variables (injection-site symptoms, n=882, 14.7% missing data; non-injection-site symptoms, n=883, 14.6% missing data). For continuous variables, the adjusted odds ratios represent the change in likelihood of side effects for a one-unit increase in the predictor variable, apart from age, where it an increase of one-unit represents an increase by decade.

The predictive power of the regression models for injection-site and non-injection-site symptoms was similar (Nagelkerke *R*^2^ of .361 and .338, respectively). The higher *R*^2^ values than in the regression models for the first dose is largely due to the marked effect of reporting of symptoms related to the first dose; before this predictor was added in the third block, the *R*^2^ values (.115 and .105) were not markedly different from those in the regression models for first-dose symptom reporting.

## Discussion

We found that 74% of people included in this study reported injection-site symptoms and 65% reported non-injection-site symptoms following their first COVID-19 vaccine dose. Following the second vaccine dose, 53% reported injection-site and 44% reported non-injection-site symptoms. The administration of vaccines as a nasal spray rather than an intramuscular injection would remove injection-site symptoms and may remove a barrier to vaccination. Clinical trials for nasal spray vaccines for COVID-19 are underway (correct at time of writing). ^25^

Clinical trial data indicate that side-effect reporting is lower for the second dose of the AstraZeneca vaccine, ^8^ which most of our participants (58%) reported receiving. Rates of commonly reported injection- and non-injection-site symptoms (fatigue, headache, fever) were within the range of those seen in clinical trial data. ^9 12 26^ Approximately 90% of symptoms reported following vaccination were attributed to the vaccine.

In line with previous research, we found that younger people were more likely to report vaccine side effects^8-10^. Women were also more likely to report symptoms, as in other studies investigating symptom reporting following COVID-19 vaccination. ^12 13^ Higher rates of symptoms are consistently perceived by females than by males in studies investigating symptom reporting. ^27 28^ Though most research points to an association between female sex and symptom perception, a recent comprehensive systematic review of factors associated with the nocebo response found little evidence for a gender effect. ^3^ We found no evidence for an association between previous or current SARS-CoV-2 infection and symptom reporting, contrary to previous research. ^12 13^ This may have been due to smaller sample sizes and wording of the item used to include current infection. Personal and clinical characteristics contributed little to the predictive strength of the regression models in this study.

Few psychological and contextual factors were significantly associated with symptom reporting following COVID-19 vaccination, except for prior symptom experience. This variable likely drove the additional predictive power (increase in Nagelkerke *R*^2^ from .116 to .361 for injection-site symptoms and from .106 to .338 for non-injection-site symptoms) when added to personal and clinical characteristics in the regression models of symptom reporting following the second vaccine dose. An important implication for policy, however, is that high uptake of the second COVID-19 vaccine dose suggests that previously experiencing symptoms from the first vaccine did not influence uptake of the second vaccine dose. One mechanism through which previous symptom experience may feed into later symptom perception is expectation. Symptom expectation is strongly associated with the nocebo effect and later symptom perception. ^3 4 29^ However, in this study, we did not find a statistically significant association between side-effect expectations at the start of the vaccine rollout in the UK and later symptom reporting. One reason for this may be that most participants were unsure whether they would experience symptoms from a COVID-19 vaccine at T1. ^17^

We investigated factors associated with side-effect expectations in our T1 survey, ^17^ as well as others theoretically associated with nocebo reporting^3^ and side-effect expectations. ^30^ However, we found few associations with symptom reporting, and in particular few associations with psychological factors. This may be due to the long period of time between measurement of psychological factors (January 2021) and symptom reporting (October 2021) and the fact that each explanatory variable was adjusted for all of the other variables in the statistical model. For example, we found no evidence for an association between perceived vaccine safety and side-effect reporting. In the UK, there was a media flurry around vaccine safety in April 2021, when news broke that the AstraZeneca vaccine may have been linked to unusual blood clots with low platelets. ^31^ This occurred after we had measured the psychological factors used as predictors in this study, and is likely to have affected perceptions. Studies have found that media reporting is associated with symptom reporting from the COVID-19 vaccine. ^14 15^ Investigation of possible associations of the influence of media and social media on symptom reporting was outside the scope of the study.

Strengths of this study include its large sample size and consequent power to detect small effects. Limitations include that our outcome measure (symptom reporting after the first and second COVID-19 vaccines) was measured in October 2021. While the second wave of data collection was timed to coincide with when all UK adults had been offered at both vaccine doses, thus avoiding systematic biases within the data, some participants may have completed their vaccine schedule some months before. Recall for symptoms can fade quickly. ^32^ Therefore symptom reporting, especially for the first vaccine dose (generally given 12 weeks before the second vaccine dose) may be affected by recall bias. We were unable to investigate factors associated with symptom attribution, as very few people did not attribute their symptoms to the vaccine (first and second vaccine dose). We were unable to investigate whether reporting side effects following the first vaccine dose affected uptake of the second dose due to small numbers. As so few people reported only having one dose (2%, *n*=25/1034), we assume there was no impact. This is supported by other research finding that experiencing side effects from the initial course of the COVID-19 vaccine (two doses) did not affect intention to receive a booster dose. ^33^ Few people reported receiving a Moderna vaccine and so confidence intervals are wider for these analyses, and no hypothesis tests were performed in respect of comparisons between the three vaccines in view of the disparity in the size of these subgroups. Our question measuring previous SARS-CoV-2 infection asked whether participants had previously had, or currently had, COVID-19. There may be some experience of adverse events from vaccination in those infected with SARS-CoV-2 when vaccinated. ^10^ Furthermore, participants may have been infected with SARS-CoV-2 after having completed the T1 questionnaire, but before receiving their first vaccine.

In conclusion, in this study, more people reported injection-site symptoms than non-injection-site symptoms. Symptoms were more likely to be reported following the first compared to the second vaccine dose. Approximately 90% of people reporting symptoms attributed them to the vaccine. Women and younger people were more likely to report symptoms, in line with clinical trial data. The factor most strongly associated with symptom reporting following the second vaccine dose was reporting symptoms from the first vaccine. However, few people had only had one vaccine, suggesting that perception of side effects did not deter people from having their second vaccine. Few psychological factors were associated with side-effect reporting, possibly due to the long time period between waves of data collection (January and October 2021).

## Data Availability

Data are available online.

https://osf.io/tehg8/

### Appendix A

**Table A.1.** Results of the logistic regression models for block one (personal and clinical characteristics only) analysing associations with symptom reporting following the first dose of a COVID-19 vaccination. For continuous variables, the adjusted odds ratios (aORs) represent the change in likelihood of side effects for a one-unit increase in the predictor variable, apart from age, where it an increase of one-unit represents an increase by decade.

|  | Predictor variable | Level | Injection-site symptoms, *n*=900 |  | Non-injection-site symptoms, *n*=901 |  |
| --- | --- | --- | --- | --- | --- | --- |
|  |  |  | Adjusted odds ratio (95% CI) for reporting symptoms † | *p* value | Adjusted odds ratio (95% CI) for reporting symptoms † | *p* value |
| Block 1 – personal and clinical characteristics | Age | Decade | 0.778 (0.687 to 0.881) | <0.001* | 0.714 (0.639 to 0.798) | <0.001* |
|  | Gender | Female | Reference | - | Reference | - |
|  |  | Male | 0.359 (0.260 to 0.495) | <0.001* | 0.665 (0.499 to 0.885) | 0.005* |
|  | Ethnicity | White | Reference | - | Reference | - |
|  |  | Other ethnic groups | 0.944 (0.563 to 1.583) | 0.827 | 1.129 (0.702 to 1.816) | 0.617 |
|  | At risk for COVID-19 | No | Reference | - | Reference | - |
|  |  | Yes | 0.963 (0.646 to 1.435) | 0.852 | 0.853 (0.591 to 1.232) | 0.398 |
|  | Think previously or currently had COVID-19 | No | Reference | - | Reference | - |
|  |  | Yes | 0.881 (0.548 to 1.418) | 0.603 | 1.242 (0.807 to 1.911) | 0.325 |
|  | Vaccine brand |  |  | 0.001* |  | <0.001* |
|  |  | Pfizer-BioNTech | Reference |  | Reference |  |
|  |  | AstraZeneca | 0.530 (0.374 to 0.752) |  | 2.073 (1.514 to 2.837) |  |
|  |  | Moderna | 2.059 (0.462 to 9.176) |  | 1.13 (0.45 to 2.85) |  |

†Adjusting for age, gender, ethnicity, being at risk for COVID-19, think or had COVID-19 previously or currently, and vaccine brand.
\* $p \leq 0.05$

**Table A.2.** Results of the logistic regression models for block one (personal and clinical characteristics only) analysing associations with symptom reporting following the second dose of a COVID-19 vaccination. For continuous variables, the adjusted odds ratios (aORs) represent the change in likelihood of side effects for a one-unit increase in the predictor variable, apart from age, where it an increase of one-unit represents an increase by decade.

|  | Predictor variable | Level | Injection-site symptoms, *n*=882 | *p* value | Non-injection-site symptoms, *n*=883 | *p* value |
| --- | --- | --- | --- | --- | --- | --- |
|  |  |  | Adjusted odds ratio (95% CI) for reporting symptoms † |  | Adjusted odds ratio (95% CI) for reporting symptoms † |  |
| Block 1 – personal and clinical characteristics | Age | Decade | 0.844 (0.760 to 0.937) | 0.001* | 0.813 (0.733 to 0.903) | <0.001* |
|  | Gender | Female | Reference | - | Reference | - |
|  |  | Male | 0.579 (0.439 to 0.764) | <0.001* | 0.666 (0.504 to 0.880) | 0.004* |
|  | Ethnicity | White | Reference | - | Reference | - |
|  |  | Black and minority ethnic | 1.219 (0.782 to 1.900) | 0.382 | 1.276 (0.826 to 1.969) | 0.272 |
|  | At risk for COVID-19 | No | Reference | - | Reference | - |
|  |  | Yes | 1.138 (0.795 to 1.628) | 0.481 | 1.395 (0.974 to 1.998) | 0.069 |
|  | Think previously or currently had COVID-19 | No | Reference | - | Reference | - |
|  |  | Yes | 1.153 (0.769 to 1.727) | 0.492 | 1.429 (0.962 to 2.121) | 0.08 |
|  | Vaccine brand |  |  | <0.001* |  | 0.002* |
|  |  | Pfizer-BioNTech | Reference |  | Reference |  |
|  |  | AstraZeneca | 0.534 (0.396 to 0.727) |  | 0.759 (0.563 to 1.025) |  |
|  |  | Moderna | 2.471 (0.809 to 7.545) |  | 6.044 (1.739 to 21.006) |  |

†Adjusting for age, gender, ethnicity, being at risk for COVID-19, think or had COVID-19 previously or currently, and vaccine brand.
\* $p \leq 0.05$

## References

1. Barsky AJ, Saintfort R, Rogers MP, et al. Nonspecific medication side effects and the nocebo phenomenon. JAMA 2002;287(5):622–7. doi: 10.1001/jama.287.5.622

2. Faasse K, Petrie KJ. The nocebo effect: patient expectations and medication side effects. Postgrad Med J 2013;89(1055):540–6. doi: 10.1136/postgradmedj-2012-131730

3. Webster RK, Weinman J, Rubin GJ. A systematic review of factors that contribute to nocebo effects. Health Psychol 2016;35(12):1334–55. doi: 10.1037/hea0000416

4. Smith LE, Weinman J, Amlot R, et al. Parental Expectation of Side Effects Following Vaccination Is Self-fulfilling: A Prospective Cohort Study. Ann Behav Med 2019;53(3):267–82. doi: 10.1093/abm/kay040

5. Faasse K, Porsius JT, Faasse J, et al. Bad news: The influence of news coverage and Google searches on Gardasil adverse event reporting. Vaccine 2017;35(49 Pt B):6872–78. doi: 10.1016/j.vaccine.2017.10.004

6. Nestoriuc Y, Orav EJ, Liang MH, et al. Prediction of nonspecific side effects in rheumatoid arthritis patients by beliefs about medicines. Arthritis Care Res (Hoboken) 2010;62(6):791–9. doi: 10.1002/acr.20160

7. Cooper V, Metcalf L, Versnel J, et al. Patient-reported side effects, concerns and adherence to corticosteroid treatment for asthma, and comparison with physician estimates of side-effect prevalence: a UK-wide, cross-sectional study. NPJ Prim Care Respir Med 2015;25:15026. doi: 10.1038/npjpcrm.2015.26

8. Voysey M, Clemens SAC, Madhi SA, et al. Safety and efficacy of the ChAdOx1 nCoV-19 vaccine (AZD1222) against SARS-CoV-2: an interim analysis of four randomised controlled trials in Brazil, South Africa, and the UK. Lancet 2021;397(10269):99–111. doi: 10.1016/S0140-6736(20)32661-1

9. Polack FP, Thomas SJ, Kitchin N, et al. Safety and Efficacy of the BNT162b2 mRNA Covid-19 Vaccine. N Engl J Med 2020;383(27):2603–15. doi: 10.1056/NEJMoa2034577

10. Baden LR, El Sahly HM, Essink B, et al. Efficacy and Safety of the mRNA-1273 SARS-CoV-2 Vaccine. N Engl J Med 2021;384(5):403–16. doi: 10.1056/NEJMoa2035389

11. Thomas SJ, Moreira ED, Jr., Kitchin N, et al. Safety and Efficacy of the BNT162b2 mRNA Covid-19 Vaccine through 6 Months. N Engl J Med 2021;385(19):1761–73. doi: 10.1056/NEJMoa2110345

12. Menni C, Klaser K, May A, et al. Vaccine side-effects and SARS-CoV-2 infection after vaccination in users of the COVID Symptom Study app in the UK: a prospective observational study. Lancet Infect Dis 2021;21(7):939–49. doi: 10.1016/S1473-3099(21)00224-3

13. Beatty AL, Peyser ND, Butcher XE, et al. Analysis of COVID-19 Vaccine Type and Adverse Effects Following Vaccination. JAMA Netw Open 2021;4(12):e2140364. doi: 10.1001/jamanetworkopen.2021.40364

14. Clemens KS, Faasse K, Tan W, et al. Social Pathways to Side-Effects: Personal Contacts and Social Media Predict COVID-19 Vaccine Side-Effect Expectations and Experience. PsyArXiv 2022. doi [preprint]: 10.31234/osf.io/e2bfv

15. MacKrill K. Impact of media coverage on side effect reports from the COVID-19 vaccine. Personal Communication.

16. Geers AL, Clemens KS, Faasse K, et al. Psychosocial Factors Predict COVID-19 Vaccine Side Effects. Psychother Psychosom 2022;91(2):136–38. doi: 10.1159/000519853

17. Smith LE, Sim J, Amlot R, et al. Side-effect expectations from COVID-19 vaccination: Findings from a nationally representative cross-sectional survey (CoVAccS - wave 2). J Psychosom Res 2021;152:110679. doi: 10.1016/j.jpsychores.2021.110679

18. Smith LE, Sim J, Cutts M, et al. Psychosocial factors affecting COVID-19 vaccine uptake in the UK: a prospective cohort study (CoVAccS – wave 3). medRxiv 2022:2022.03.25.22272954. doi [pre-print]: 10.1101/2022.03.25.22272954

19. Sherman SM, Smith LE, Sim J, et al. COVID-19 vaccination intention in the UK: results from the COVID-19 vaccination acceptability study (CoVAccS), a nationally representative cross-sectional survey. Hum Vaccin Immunother 2021;17(6):1612–21. doi: 10.1080/21645515.2020.1846397

20. Open Science Framework. COVID-19 vaccination acceptability in the UK January 2021 (CoVAccS-Wave 2) 2021 [updated 14 October 2021]. Available from: https://osf.io/ewch3/ (Accessed 25 July 2022).

21. Open Science Framework. COVID-19 vaccination-factors affecting uptake in the UK (CoVAccS wave 3) 2022 [updated 18 February 2022]. Available from: https://osf.io/tehg8/ (accessed 1 March 2022).

22. MacKrill K, Webster R, Rubin GJ, et al. When symptoms become side effects: Development of the side effect attribution scale (SEAS). J Psychosom Res 2021;141:110340. doi: 10.1016/j.jpsychores.2020.110340

23. Sherman SM, Sim J, Cutts M, et al. COVID-19 vaccination acceptability in the UK at the start of the vaccination programme: a nationally representative cross-sectional survey (CoVAccS-wave 2). Public Health 2022;202:1–9. doi: 10.1016/j.puhe.2021.10.008

24. Chen H, Cohen P, Chen S. How Big is a Big Odds Ratio? Interpreting the Magnitudes of Odds Ratios in Epidemiological Studies. Communications in Statistics - Simulation and Computation 2010;39(4):860–64. doi: 10.1080/03610911003650383

25. Mahase E. Sixty seconds on nasal vaccines. BMJ 2022;377:o1148. doi: 10.1136/bmj.o1148

26. Folegatti PM, Ewer KJ, Aley PK, et al. Safety and immunogenicity of the ChAdOx1 nCoV-19 vaccine against SARS-CoV-2: a preliminary report of a phase 1/2, single-blind, randomised controlled trial. Lancet 2020;396(10249):467-78. doi 10.1016/S0140-6736(20)31604-4

27. van Wijk CM, Kolk AM. Sex differences in physical symptoms: the contribution of symptom perception theory. Soc Sci Med 1997;45(2):231–46. doi: 10.1016/s0277-9536(96)00340-1

28. Barsky AJ, Peekna HM, Borus JF. Somatic symptom reporting in women and men. J Gen Intern Med 2001;16(4):266–75

29. Rheker J, Winkler A, Doering BK, et al. Learning to experience side effects after antidepressant intake - Results from a randomized, controlled, double-blind study. Psychopharmacology (Berl) 2017;234(3):329–38. doi: 10.1007/s00213-016-4466-8

30. Smith LE, Webster RK, Rubin GJ. A systematic review of factors associated with side-effect expectations from medical interventions. Health Expect 2020;23(4):731–58. doi: 10.1111/hex.13059

31. European Medicines Agency. AstraZeneca’s COVID-19 vaccine: EMA finds possible link to very rare cases of unusual blood clots with low blood platelets 2021 [updated 7 April 2021]. Available from: https://www.ema.europa.eu/en/news/astrazenecas-covid-19-vaccine-ema-finds-possible-link-very-rare-cases-unusual-blood-clots-low-blood (accessed 1 October 2021).

32. Feikin DR, Audi A, Olack B, et al. Evaluation of the optimal recall period for disease symptoms in home-based morbidity surveillance in rural and urban Kenya. Int J Epidemiol 2010;39(2):450–8. doi: 10.1093/ije/dyp374

33. Geers AL, Clemens KS, Colagiuri B, et al. Do Side Effects to the Primary COVID-19 Vaccine Reduce Intentions for a COVID-19 Vaccine Booster? Ann Behav Med 2022 doi: 10.1093/abm/kaac027

